# Impact of Omicron variant on the response to SARS-CoV-2 mRNA Vaccination in multiple myeloma

**DOI:** 10.1101/2022.02.25.22271501

**Authors:** Paola Storti, Valentina Marchica, Rosanna Vescovini, Valentina Franceschi, Luca Russo, Vincenzo Raimondi, Denise Toscani, Jessica Burroughs Garcia, Federica Costa, Benedetta Dalla Palma, Naomi Soressi, Mariateresa Giaimo, Nicolas Thomas Iannozzi, Laura Notarfranchi, Gabriella Sammarelli, Gaetano Donofrio, Nicola Giuliani

## Abstract

Multiple myeloma (MM) patients may have a reduced response to vaccination due to immunodeficiency. The humoral and cellular response to SARS-CoV-2 mRNA full vaccination and booster dose as well as the impact of spike variants, including the emerging Omicron one, are still unclear and have been investigated in this study in a cohort of MM patients and those with pre-malignant monoclonal gammopathies.

Firstly, we have shown that MM patients with relapsed-refractory disease (MMR) had a reduced spike-specific antibody levels and neutralizing titers after SARS-CoV-2 mRNA full vaccination. Interestingly, all the analyzed variants, remarkably Omicron, had a significant negative impact on the neutralizing ability of the vaccine-induced antibodies in all patients with MM and in smoldering MM too. Moreover, lower spike-specific IL-2-producing CD4^+^ T cells and reduced cytotoxic spike-specific IFN-γ and TNF-α-producing-CD8^+^ T cells were found in MM patients as compared to MGUS.

Finally, we found that booster immunization improved SARS-CoV-2 spike humoral and cellular responses in newly diagnosed MM (MMD) patients and in most, but not all, MMR patients. After the booster dose, a significant increase of the neutralizing antibody titers against almost all the analyzed variants was achieved in MMD. On the other hand, in MMR patients, Omicron retain a negative impact on neutralizing ability, suggesting these patients need to be considered still at risk of Omicron SARS-CoV-2 infection with a clinically relevant disease.

## Introduction

Multiple myeloma (MM) is a hematological malignancy characterized by impairment of both cellular and humoral responses,^1^ with high risk of infection including viral and bacterial ones.^2^ Alterations of immune response to infections have been also describe in the pre-malignant state of monoclonal gammopathies as monoclonal gammopathies of undetermined significance (MGUS) and smoldering myeloma (SMM); although the risk of infections in this type of patients is lower as compared to MM patients.^2–4^ The well-known severe acute respiratory syndrome coronavirus 2 (SARS-CoV-2) pandemic infection completely changed the epidemiological scenario also in hematological patients including MM due to either their intrinsic immunological defects or to the effect of the immune-suppressive treatment. It has been reported that COVID-19 causes moderate to severe acute respiratory disease in approximately 75% to 80% of patients with MM, resulting in death in almost one-third of hospitalized patients.^5,6^ Thus, vaccination against SARS-CoV-2 is the most important preventive strategy to protect MM patients from COVID-19. Published data show that MM patients share a reduced antibody production in response to anti-SARS-CoV-2 vaccination.^7,8^ In addition, MM patients treated with anti-CD38 or anti–B-cell maturation antigen (BCMA) antibodies, seem not to develop anti–SARS-CoV-2 antibodies or have insufficient response even after full vaccination.^9,10^ However, very few data are actually available about the cellular response to SARS-CoV-2 mRNA vaccination^11^ and the efficacy of the mRNA vaccine, including the booster dose, to the different SARS-CoV-2 variant characterized by mutations encoding for the spike protein.^12^

Since November 2021, the B.1.1.529 (Omicron) variant has rapidly become dominant globally.^13^ Omicron partially evades antibodies induced by infection or vaccination and it raises concerns regarding the effectiveness of current vaccines.^14–16^ Few data are available on the response after full vaccination and booster dose in cancer patients^17^ and any detailed studies are available in patients with MM and monoclonal gammopathies.

In this study, we investigated both the humoral and cellular response to SARS-CoV-2 mRNA full vaccination and to the booster dose in a cohort of patients with MGUS, SMM and MM. In these patients, we quantified SARS-CoV-2 spike IgG antibodies (spike-IgG-Abs), neutralizing antibody (NAb) titers against vaccine homologous spike and five variants of concern (VoC), including Omicron, and SARS-CoV-2 spike-specific CD4^+^ and CD8^+^ T cells. Moreover, in MM patients we also studied the effect of a booster dose on the SARS-CoV-2 spike-specific immune responses, especially, on the neutralizing capacity against the spike variants.

## Methods

### Study information and patient clinical characteristic

All patients were followed and treated at the Hematology Unit of Parma Hospital and received vaccination as part of the national COVID-19 vaccination program. From February 25 to July 23, 2021, 40 patients were enrolled in the study: 6 MGUS, 10 SMM and 24 MM patients, including either newly diagnosed MM (MMD) or relapsed-refractory MM (MMR). All MMD are in first line of treatment whereas MMR underwent to at least two lines of treatment.

Peripheral blood (PB) samples were collected at two time points: before the first dose (PRE) and 14±2 days after the second dose (POST), i.e 35 days after the first dose of the BNT162b2, mRNA vaccine by Pfizer-BioNTech (Fig 1A). In a subset of 16 patients with MM (MMD and MMR), blood samples were also collected after 14±2 days of a heterologous booster dose (BOOSTER) with mRNA-1273 by Moderna, received, in November 2021, at least six months (>180 days) after the complete vaccination (Fig.4A)

**Fig 1.**
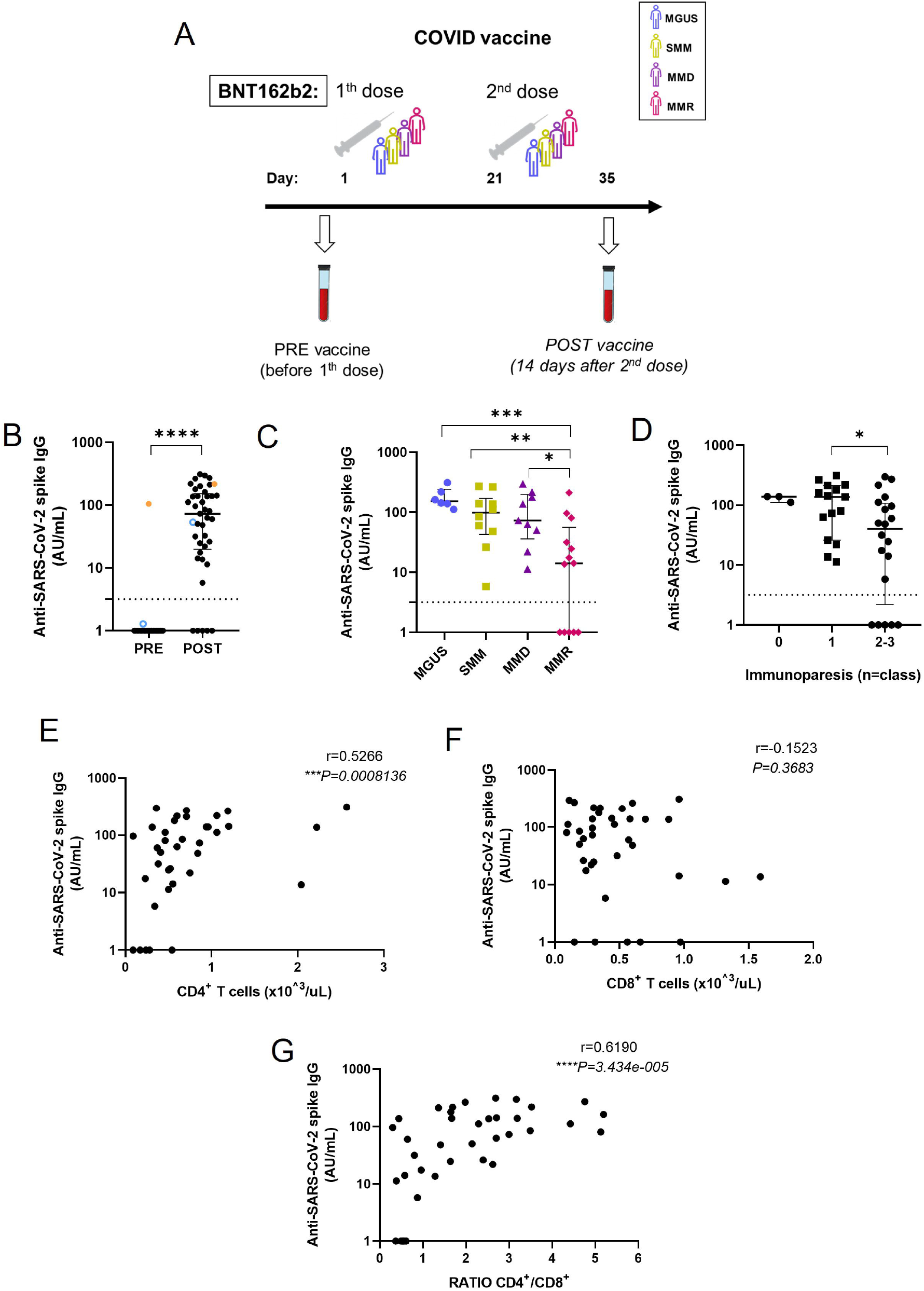
Vaccine-induced SARS-CoV-2 spike IgG antibodies in patients with monoclonal gammopathies at different stages of disease. **(A)** Study design, vaccine administration scheme and time points collected for patients. **(B)** SARS-CoV-2 spike IgG antibodies were measured PRE and POST vaccination (n = 40 patients), using the COVID-SeroIndex Kantaro SARS-CoV-2 IgG test. Quantitative results are reported in Arbitrary Units/ml (AU/mL) with the lower limit of detection at <3.20 (AU/mL). Individual data points are shown as scatter dot plot with lines showing the median with IQR. Dotted lines show the lower limit of detection. Orange dots identify a patient with highly suspected pre-vaccination COVID-19 infection and light blue dots a patient with a PCR-proven COVID-19 infection between the two time points. Statistical analysis was performed using a paired, two-tailed, nonparametric Wilcoxon test. (**C)** SARS-CoV-2 spike IgG antibody levels in patients subdivided in the different stages of disease: MGUS (indigo ●) (n =6); SMM (avocado ■) (n =10); MMD (purple ▲) (n = 9) and MMR (fuchsia ♦) (n = 13). Significant difference was determined by two-tailed Mann–Whitney U tests. **(D)** SARS-CoV-2 spike IgG antibody levels in relation to number of IgG classes involved in immunoparesis at the time of vaccination. **(E)** Correlations between levels of SARS-CoV-2 spike IgG antibodies after vaccination and the baseline (PRE) percentage of total CD4+ T cells or (**F**) percentage of total CD8+ T cells or (g) ratio CD4+/CD8+. Correlations were calculated using nonparametric Spearman rank correlation. *P* values are shown when *P* < 0.05 (\**P* < 0.05, \*\**P* < 0.01, \*\*\**P* < 0.001)

### Ethics statement

PB samples were obtained according to the criteria of the Declaration of Helsinki and following written informed consent. The study was approved by the Local Ethics Committee.

### Detection of SARS-CoV-2-specific IgG antibodies

Heat inactivated sera samples were tested for SARS-CoV-2-specific IgG antibodies using a commercial quantitative two-step ELISA (COVID-SeroIndex, Kantaro Quantitative SARS-CoV-2 IgG Antibody Kit, R&D Systems), according to the manufacturer’s recommendations. Quantitative results are reported in Arbitrary Units/ml (AU/mL) with the lower limit of detection at <3.20 (AU/mL).

### SARS-CoV-2 pseudoviruses generation and neutralization assay against the original viral strain and variants

Lentiviral vector-based SARS-CoV-2 spike pseudoviruses were generated as previously described ^18^ with minor modifications as described in Supplemental Data. SARS-CoV-2 spike pseudoviruses displayed on their surface 6 different spike glycoproteins: Wuhan-Hu-1 (B.1 Lineage; China) Alpha (B.1.1.7. Lineage; United Kingdom), Beta (B.1.351 Lineage; South Africa), Gamma (P.1 Lineage; Brasil), Delta (B.1.617.2 Lineage, India) or Omicron (B.1.1.529 Lineage; Europe).

The neutralization assay was performed on 104 HEK/ACE2/TMPRRS2/Puro cells ^18^ testing heat inactivated sera samples at the dilution of 1:4–1:8–1:16–1:32–1:64–1:128–1:256–1:512 as described in Supplemental Methods. A negative control was established without serum. The RLUs were compared and normalized to those derived from wells where pseudovirus was added in the absence of sera (100%). Neutralization titer 50 (NT50) was expressed as the maximal dilution of the sera where the reduction of the signal is ≥50%. Each serum was tested in triplicate.

### Intracellular Cytokine Staining Flow Cytometry (ICS) T Cell Assay

Patients’ PBMCs were thawed, resuspended in RPMI media supplemented with 10% heat-inactivated fetal calf serum (Biochrom, GmbH), 2 mM L-glutamine and 1% penicillin–streptomycin (R10) and rested at 37 °C for 6h. After resting, 1×10^6^ cells of each sample were supplemented with R10 containing CD107a (cat. 555801, BD Pharmingen), monensin (cat.554724, BD Golgi Stop, BD Biosciences) and S1 and S2 peptide pools (PepMIX SARS-CoV-2 spikeglicoprotein, cat. PM-WCPV-S-3, JPT Peptide Techonologies GmbH) added at a final concentration of 1 μg/ml. For each tested sample, a positive control (*S. enterotoxin B* at 2 μg/ml, Sigma Aldrich) and an unstimulated control (stimulation with an equimolar amount of DMSO) were also included. Cells were incubated for 2h at 37 °C with 5% CO2 and then R10 containing brefeldin A (5 μg ml^−1^, Sigma Aldrich) were added and the samples were incubated for 16 h. PBMCs were washed and stained with BD Horizon Fixable Viability stain 575V (1:1000). A surface staining cocktail was added containing saturating concentrations of BV480 CD3 (cat.566105, BD Horizon), BV786 CD4 (cat.563877, BD Horizon) and BV711 CD8 (cat.563677, BD Horizon). PBMCs were fixed and permeabilized with FACS Lysins Solution 1x (cat.349202, BD Bioscience) and FACS Permeabilizing Solution2 1x (cat. 340973, BD Biosciences). After washes, PBMCs were stained with a cocktail of anti-human IFN-γ-FITC (cat.554700, BD Pharmingen), IL-2-PerCP-Cy5.5 (cat.560708, BD Pharmingen), and TNF-α-BV421 (cat.562783, BD Horizon). Samples were acquired on a BD Bioscience FACSCelesta flow cytometer using the FACSDiva Software (version 8.02, BD Bioscience). A hierarchical gating strategy was created during assay qualification and was applied for all PRE, POST and BOOSTER vaccine sample analysis (Suppl. Fig. 1 A,B). Peptide-specific responses were calculated by subtraction of the unstimulated controls from the paired peptide-stimulated samples, then, POST vaccination and BOOSTER responses were subtracted of the paired PRE vaccination responses. Negative values were designated as zero and data are represented as a percentage of total CD4+ or CD8+ T cells with the lower limit of detection at <0.01 of the parental gate.

### Statistical Analysis

Data are presented as medians with IQRs. Data analysis, as well as all graphical representation of the data, were performed in GraphPad Prism v.8.0.1 software. \**P* < 0.05, \*\**P* < 0.01, \*\*\**P* < 0.001, \*\*\*\**P* < 0.0001. (Supp. Methods)

### Data Sharing Statement

For original data, please contact.

## Results

### Antibody responses to vaccination in patients with MM and pre-malignant monoclonal gammopathies

We evaluated spike-IgG-Abs PRE and POST BNT162b2 mRNA vaccination in 40 patients with monoclonal gammopathies at different stages of disease. (Fig. 1A, B). Two patient were excluded, one because pre-existing antibodies (Fig 1B, orange dots) and the second because having COVID-19 infection between the time points. (Fig 1B, light blue circles). Thus, we focused on 38 COVID-naïve patients: 6 low-intermediate risk MGUS, 10 SMM, 9 MMD and 13 MMR, described in Supplementary Table 1.

The seropositivity rate for Spike-IgG-Abs in the total cohort was 86.8% (n. 33) with 5 (13.2%) patients exhibiting no detectable antibodies (Fig.1B). We did not find any significant correlation of the humoral response with patients’ age (Suppl. Fig. 2A). Looking at the different stages of disease, we found that MMR patients had significantly lower spike-IgG-Abs as compared with MGUS, SMM and MMD patients (Fig. 1C).

Moreover, we found that all 5 seronegative patients had immunoparesis in 2/3 Ig classes and that, overall, the presence of immunoparesis in 2/3 classes was associated with lower spike-IgG-Abs compared to involvement of only 1 class of Ig (Fig. 1D).

Considering other immune parameters possible affecting the vaccine-induced humoral responses, we did not find any significant correlation between spike-IgG-Abs responses and the baseline (PRE) total lymphocyte counts although they were significantly reduced in MMR and MMD patients compared with MGUS patients (Suppl. Fig. 2B,C). Interestingly, there was a significant correlation between spike-IgG-Abs and the baseline total CD4^+^ T cells counts (Fig. 1E), but not with total CD8^+^ T cells counts (Fig. 1F) and with the baseline CD4^+^/CD8^+^ ratio (Fig. 1G). In line with literature data, we found a significantly reduced CD4^+^/CD8^+^ ratio in MMR and also in MMD compared with the other monoclonal gammopathies. (Suppl. Fig. 2D).

To examine the functional quality of vaccine-induced antibodies, we tested patients’ sera using a neutralization assay with a pseudovirus displaying on its surface vaccine homologous spike (Wuhan-Hu-1 sequence). Looking at the different stages of disease, we found that MMR patients had significantly lower NAb titers compared with MGUS, SMM and MMD patients (Fig. 2A). Notably, 5 out of 6 patients with a neutralizing titer below the detection level were MMR patients. We found a significant correlation between spike-IgG-Abs levels and neutralizing titers against Wuhan-Hu-1 spike (Fig. 2B).

**Fig 2.**
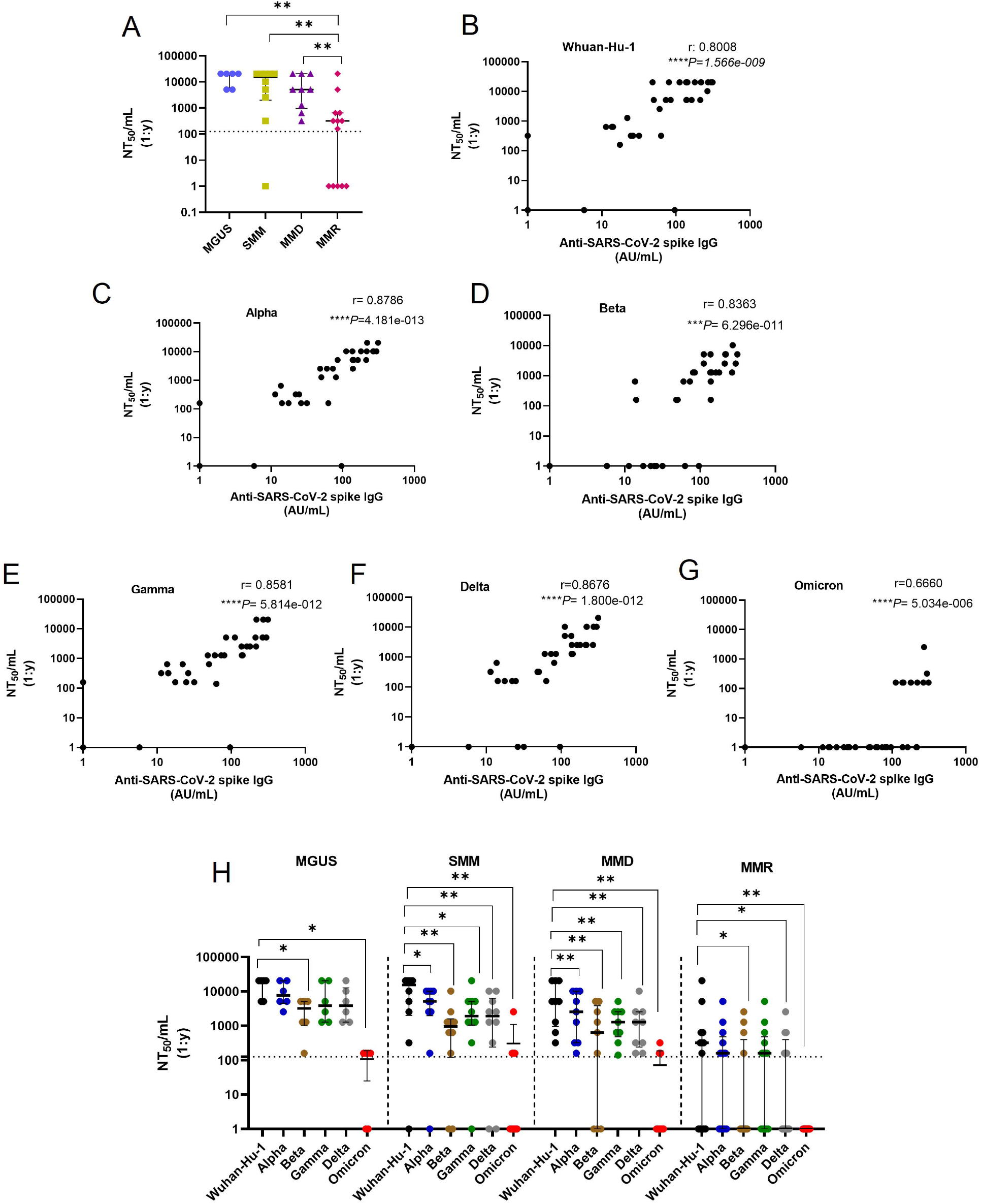
Vaccine-induced SARS-CoV-2 spike neutralizing antibody titers in patients with monoclonal gammopathies at different stages of disease. Neutralizing antibody titers were measured POST vaccination using a neutralization assay with pseudoviruses expressing the Whuan-Hu-1 original spike protein and Alpha, Beta, Gamma, Delta or Omicron variants. Quantitative results are reported in Neutralization titer 50/mL (NT50/mL) as the maximal dilution of the sera where the reduction of the signal is ≥50%, with the lower limit of detection at 160 Nt50/mL. **(A)** Neutralizing antibody titers against the Whuan-Hu-1 original spike protein. Individual data points are shown as scatter dot plot with lines showing the median with IQR. Dotted lines show the lower limit of detection. Patients are subdivided in relation to the different stages of disease MGUS (indigo ●) (n =6); SMM (avocado ■) (n =10); MMD (purple ▲) (n = 9) and MMR (fuchsia ♦) (n = 13). Significant difference was determined by two-tailed Mann–Whitney U tests. **(B)** Correlations between levels of SARS-CoV-2 spike IgG antibodies and neutralizing antibody titers to Whuam-Hu-1 original spike protein or (**C**) neutralizing antibody titers to Alpha variant or (**D**) neutralizing antibody titers to Beta variant or (**E**) neutralizing antibody titers to Gamma variant, (**F**) neutralizing antibody titers to Delta variant or (**G**) neutralizing antibody titers to Omicron variant. Correlations were calculated using nonparametric Spearman rank correlation. **(H)** Comparison between neutralizing titers to Whuan-Hu-1 original spike protein and Alpha, Beta, Gamma, Delta and Omicron variants, in patients stratified for stages of disease. Individual data points are shown as scatter dot plot with lines showing the median with IQR. Dotted lines show the lower limit of detection. Statistical analysis was performed using a paired, two-tailed, nonparametric Wilcoxon test. *P* values are shown when *P* < 0.05 (\**P* < 0.05, \*\**P* < 0.01).

We next wanted to determine whether the vaccine-induced antibodies protect patients against VoC of the virus that raise concerns regarding the effectiveness of current vaccines.^19–22^ We tested patients’ sera samples in the neutralization assay with pseudoviruses displaying Alpha, Beta, Gamma, Delta or the most recent Omicron variants of the spike protein. We found significant correlations between levels of spike-IgG-Abs and NAb titers against the five variants (Fig. 2C-F), indicating that vaccine-induced antibodies retained functional characteristics and neutralizing ability against the studied variants, even if we noticed a general reduction in neutralizing titers to variants. (Fig. 2C-G).

We analyzed neutralizing titers to the five variants in comparison to Wuhan-Hu-1 spike, in patients stratified for disease stages (Fig. 2H). Interestingly, among MGUS patients, we found that NAb titers to Beta and Omicron variants were significantly lower than Wuhan-Hu-1 spike. On the contrary, among SMM and MMD patients, NAb titers to all variants were significantly lower than original spike. (Fig. 2H) In MMR patients, already showing a significant reduction in NAb titers to the Wuhan-Hu-1 spike compared with other patients (Fig. 2A), we found lower NAb titers to Beta, Delta and Omicron variants than original spike (Fig. 2H).

In particular, the neutralization ability against Omicron variant was dramatically reduced in all groups of patients (red dots in Fig.2H): in MMR patients we did not find any detectable neutralizing titer (0/13) against this variant compared to 4/6 (66.6%), 4/10 (40%), and 3/9 (33.3) in MGUS, SMM and MMD patients, respectively.

These data overall indicated that, after full vaccination, all the analyzed variants, remarkably the Omicron one, had a significant negative impact on the neutralizing ability of the vaccine-induced antibodies in SMM, MMD and MMR.

### Cellular responses to vaccination in patients with MM and pre-malignant monoclonal gammopathies

Clinical studies have suggested a protective role for both humoral and cell-mediated immunity in recovery from SARS-CoV-2 infection^23–25^ and the goal of COVID-vaccines is to elicit a coordinated immunological memory including both NAbs and SARS-CoV-2 spike-specific T cells.^26^ Today, the size and the quality of T cell responses induced by SARS-CoV-2 vaccination in patients with monoclonal gammopathies are not fully understood. Then, we evaluated vaccine-induced circulating SARS-CoV-2 spike-specific T cells by ICS flow cytometry analysis.

First, we noticed that CD4^+^ and CD8^+^ T cell responses to SARS-CoV-2 vaccine were highly variable in our cohort (Fig 3). We observed that 100% of MGUS patients had a SARS-CoV-2 spike-specific CD4^+^ T cell response (at least one cytokine production), compared to 70% of SMM, 77.7 % of MMD and 58.3% of MMR patients (Fig. 3A). In particular, we found that SMM, MMD and MMR had significantly reduced IL-2^+^CD4^+^ T cells compared to MGUS patients, while we did not find any significant differences for IFN-γ- and TNF-α producing CD4^+^ T cells (Fig 3B). Focusing on CD4^+^ T cells double positive, we found that MMR had significantly reduced IL-2+TNF-α+ CD4^+^ T cells compared to MGUS patients (Fig 3C).

**Fig. 3.**
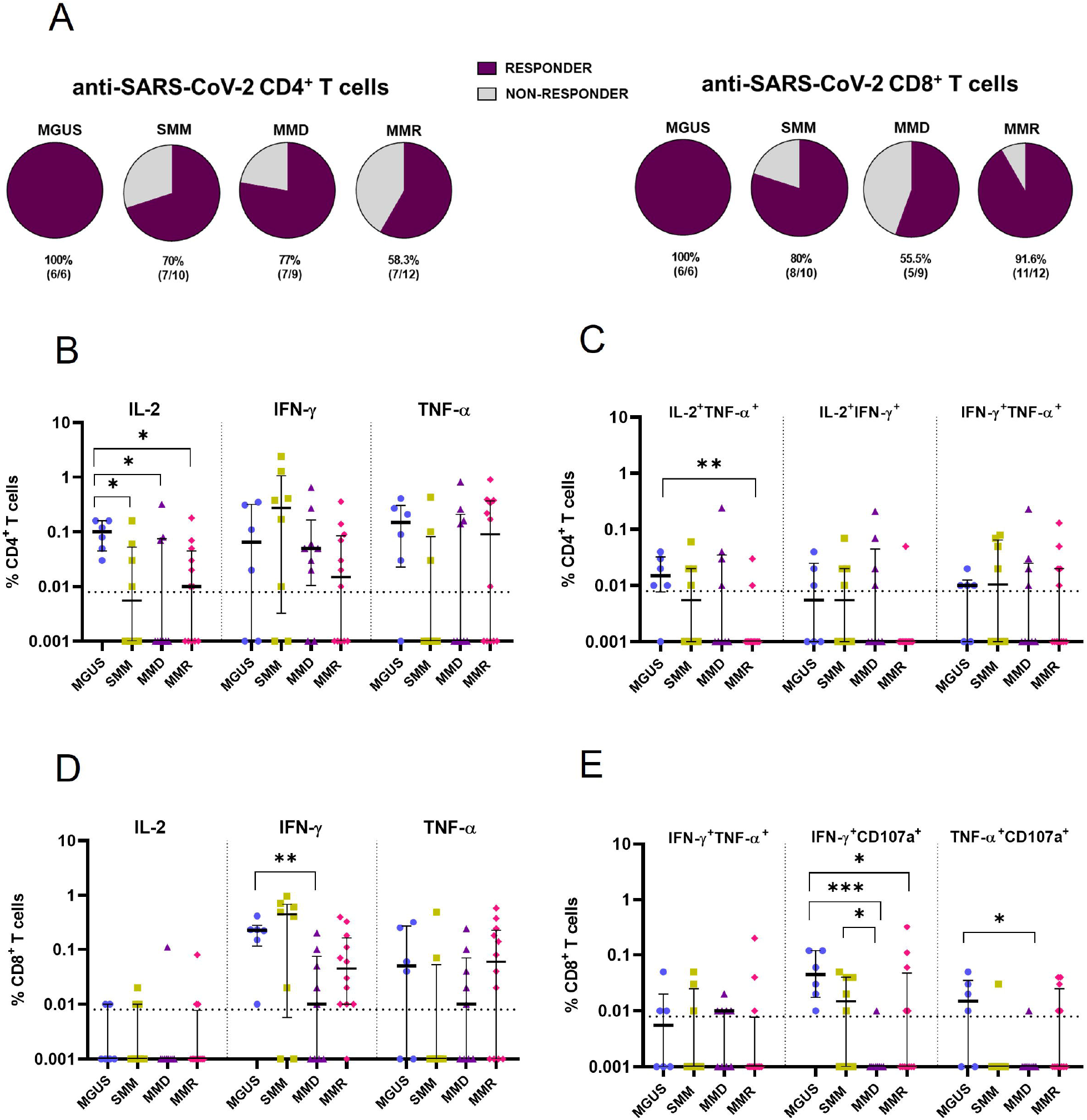
Vaccine-induced SARS-CoV-2 spike-specific T cell responses in patients with monoclonal gammopathies at different stages of disease. Circulating SARS-CoV-2 spike-specific T cells were evaluated by ICS flow cytometry analysis, after overnight stimulation of PBMCs collected PRE and POST vaccination with peptide pools covering the SARS-CoV-2 spike protein (original Wuhan-Hu-1 sequence). Peptide-specific responses were calculated by subtraction of the unstimulated controls from the paired peptide-stimulated samples and subsequently POST vaccination responses were subtracted of the paired PRE vaccination responses. Negative values were designated as zero. Data are represented as a percentage of total CD4+ or CD8+ T cells with the lower limit of detection at <0.01. **(A)** Pie graphs indicate the frequency of patients with (RESPONDER) or without (NON RESPONDER) detectable SARS-CoV-2 spike-specific CD4+ or CD8+ T cell responses, according to the different stages of disease (MGUS n=6, SMM n=10, MMD n=9 and MMR n=12). **(B)** Percentage of SARS-CoV-2 spike-specific CD4+ T cells expressing IL-2, IFN-γ, or TNF-α and (**C**) percentage of double positive CD4+ T cells for the indicated cytokines in patients at different stages of disease. **(D)** Percentage of SARS-CoV-2 spike-specific CD8+ T cells expressing IL-2, IFN-γ, or TNF-α and (**E**) percentage of double positive CD8+ T cells for the indicated cytokines or the degranulation marker CD107a in patients at different stages of disease. Individual data points are shown as scatter dot plot (MGUS (indigo ●); SMM (avocado ■); MMD (purple ▲) and MMR (fuchsia ♦)) with lines showing the median with IQR. Dotted lines show the lower limit of detection. Significant difference was determined by two-tailed Mann–Whitney U tests with *P* values shown when *P* < 0.05 (\**P* < 0.05, \*\**P* < 0.01, \*\*\**P* < 0.001).

Moreover, the percentages of patients with a SARS-CoV-2 spike-specific CD8^+^ T cell response were 100%, 80%, 55.5% and 91.6% among MGUS, SMM, MMD and MMR, respectively (Fig. 3A). In particular, we found that MMD patients had a significant reduction in IFN-γ^+^ CD8^+^ T cells compared to MGUS but any significant difference for TNF-α production (Fig 3D). Focusing on CD8^+^ T cells double positive, we found that both MMD and MMR patients had significantly reduced IFN-γ^+^CD107a^+^ CD8+ T cell responses compared to MGUS and MMD compared also to SMM patients (Fig. 3E). Moreover, MMD patients had significantly reduced TNF-α^+^CD107a^+^ CD8^+^ T cells compared to MGUS (Fig.3E).

Finally, we did not find a significant correlation between levels of spike-IgG-Abs and SARS-CoV-2-specific CD4+ or CD8+ T cells (Suppl. Fig. 3).

Taken together, these data indicate that not only MM but also SMM patients showed a reduced cellular response to SARS-CoV-2 full vaccination compared to MGUS and a reduced percentage of antigen-specific IL-2^+^CD4^+^ T cells. MM patients, concomitantly, showed a reduced cytotoxic function exerted by the IFN-γ^+^ and TNF-α^+^CD8^+^ T cells, compared to MGUS.

### Effects of the booster dose on spike-specific immune responses in patients with MM

In 16 MM patients of our cohort, 7 MMD and 9 MMR, we also evaluated the humoral and cellular spike-specific responses after a heterologous booster mRNA-1273 vaccine (Fig. 4A), Firstly, MMR patients retained significantly lower anti-spike-IgG-Abs levels compared to MMD patients, even after the booster dose (Fig 4B).

**Fig. 4.**
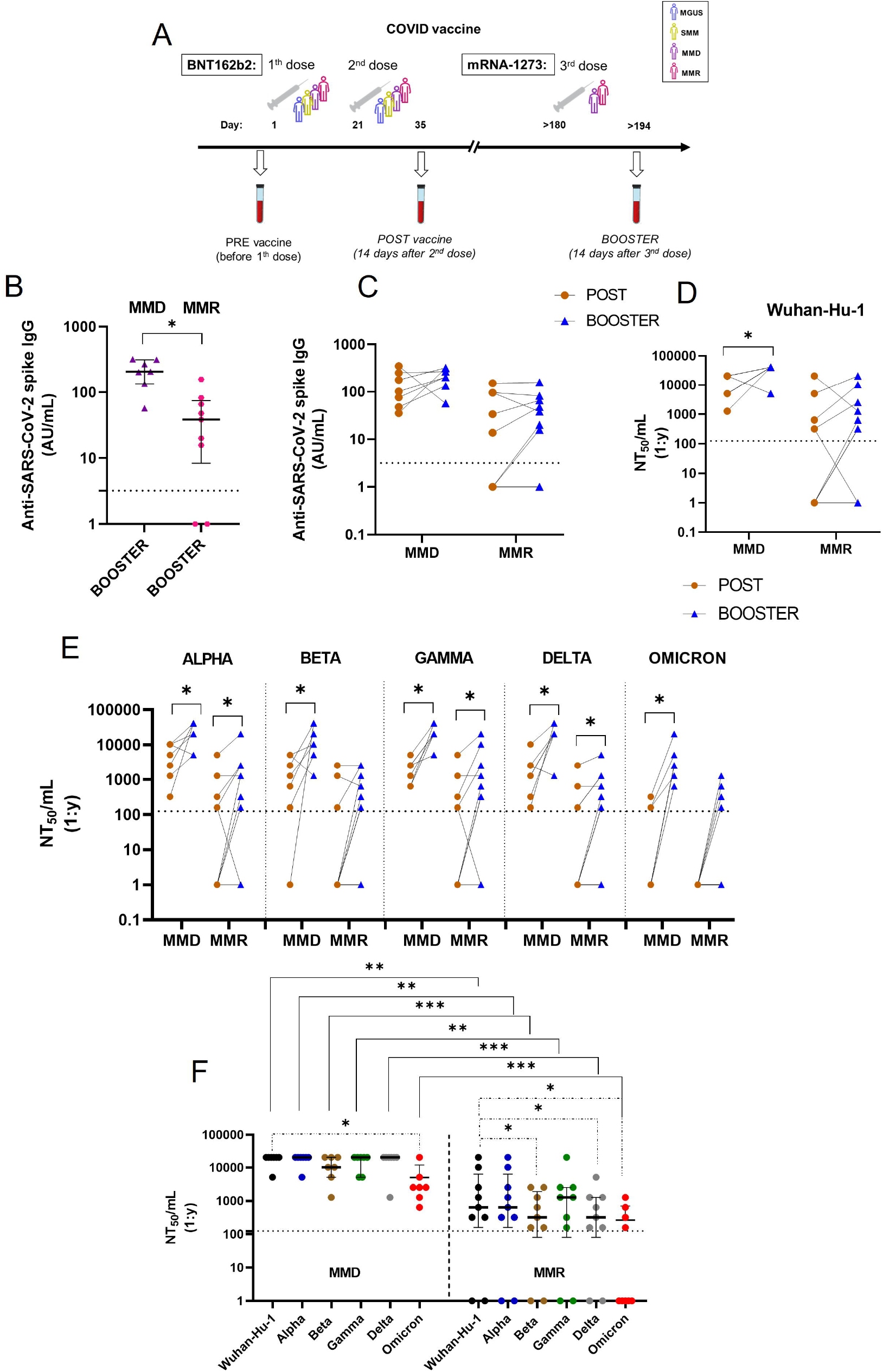
Effects of the booster dose on spike-specific humoral responses in patients with MM. **(A)** Study design, vaccine administration scheme and time points collected for patients. **(B)** SARS-CoV-2 spike IgG antibodies were measured after the third vaccine dose (BOOSTER) in 16 patients (MMD = n. 7 and MMR = n.9). Individual data points are shown as scatter dot plot with lines showing the median with IQR. Dotted lines show the lower limit of detection. Significant difference was determined by two-tailed Mann–Whitney U tests. *P* values are shown when *P* < 0.05. **(C)** SARS-CoV-2 spike IgG antibodies measured after POST (orange ●) and BOOSTER (blue ▲) in MMD and MMR patients: individual data points are shown, and lines connect the paired samples. Dotted lines show the lower limit of detection. Statistical analysis was performed using a paired, two-tailed, nonparametric Wilcoxon test. **(D)** Neutralizing antibody titers were measured in the POST (orange ●) and BOOSTER (blue ▲) samples using a neutralization assay with pseudoviruses expressing the Whuan-Hu-1 original spike protein and **(E)** Alpha, Beta, Gamma, Delta and Omicron spike protein variants. Quantitative results are reported as individual dot and lines connect paired samples and the y axis reports Neutralization titer 50/mL (NT50/mL) as the maximal dilution of the sera where the reduction of the signal is ≥50%, with the lower limit of detection at 160 NT50/mL. Dotted lines show the lower limit of detection. Statistical analysis was performed using a paired, two-tailed, nonparametric Wilcoxon test. *P* values shown when *P* < 0.05 (\**P* < 0.05). **(F)** Comparison between neutralizing titers to Whuan-Hu-1 original spike protein and Alpha, Beta, Gamma, Delta and Omicron variants, within MMD and MMR groups. Individual data points are shown as scatter dot plot with lines showing the median with IQR. Dotted lines show the lower limit of detection. Statistical analysis was performed using a paired, two-tailed, nonparametric Wilcoxon test. *P* values are shown when *P* < 0.05 (\**P* < 0.05, \*\**P* < 0.01, \*\*\**P* < 0.001).

Secondly, comparing spike-IgG-Abs levels between POST and BOOSTER, we did not find a significant increase both in MMD and MMR patients. However, after the booster dose, 85.7% (6/7) of MMD patients improved spike-IgG-Abs levels and among MMR patients, 50% (2/4) of the seronegative patients, in the POST sample, had a seroconversion and 60% of the seropositive patients (3/5) had an increase in spike-IgG-Abs levels. (Fig. 4C)

Subsequently, we found a significant increase in the NAb titer to the Whuan-Hu-1 spike in MMD but not in MMR patients, although 6/9 (66.6%) MMR patients improved their titers respect the POST sample. (Fig.4D).

Interestingly, the NAb titers against spike variants, including the Omicron one, significantly increased after the booster compared to the second dose in MMD patients (Fig. 4E). Among MMR patients, we found a significant increase in the NAb titer to the Alpha, Gamma and Delta variants, but not to Beta and Omicron variants. (Fig. 4E). However, 4/9 (44.4%) of MMR patients reached delectable levels of NAbs against Omicron variant in BOOSTER compared to POST samples, where this titer was undetectable in 100% of the samples.

Despite this last result, MMR patients retained significantly lower titers compared to MMD patients against the Whuan-Hu-1 spike and the five variants, even after the booster dose. (Fig. 4F, solid comparison lines)

Interestingly, after the booster dose, MMD patients lost the negative impact of the spike variants seen with the full vaccination. In fact, we did not observe any significant difference compared to Wuhan-Hu-1 spike in NAb titers to four variants, except for Omicron (Fig. 4F dotted comparison line); nevertheless, all MMD patients reached detectable levels of NAbs against Omicron. MMR patients retained NAb titers to Beta, Delta and Omicron variants significantly lower than Whuan-Hu-1 spike. (Fig. 4F, dotted comparison lines)

Finally, we explored the impact of the heterologous BOOSTER on the cellular responses. We noticed that CD4^+^ and CD8^+^ T cell responses to SARS-CoV-2 vaccine were highly variable in our cohort still after the booster dose (Fig. 5).

**Fig. 5.**
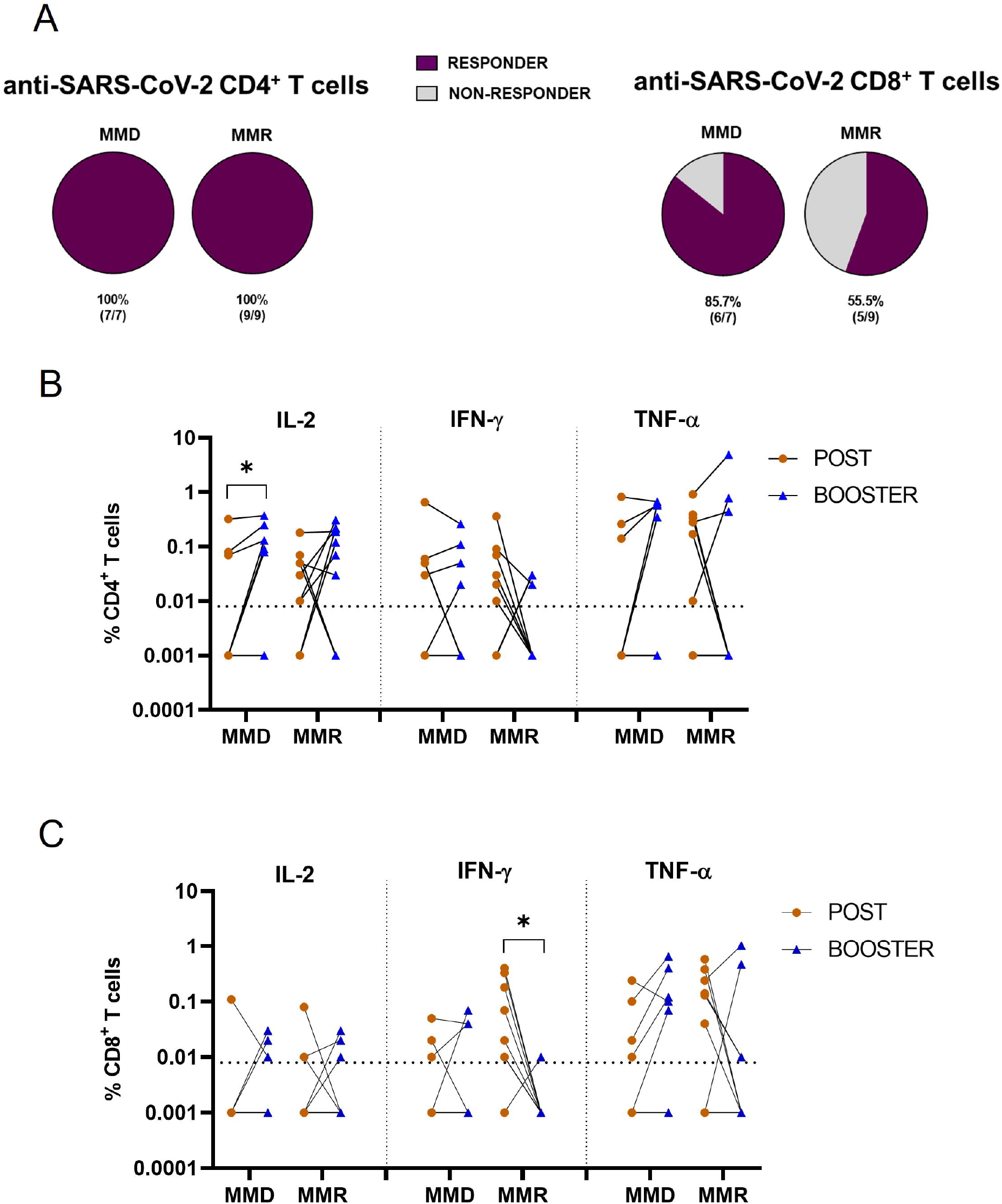
Effects of the booster dose on spike-specific cellular responses in patients with MM. Circulating SARS-CoV-2 spike-specific T cells were evaluated by ICS flow cytometry analysis, after overnight stimulation of PBMCs with peptide pools covering the SARS-CoV-2 spike protein (original Wuhan-Hu-1 sequence). Peptide-specific responses were calculated by subtraction of the unstimulated controls from the paired peptide-stimulated samples and subsequently POST vaccination and BOOSTER responses were subtracted of the paired PRE vaccination responses. Negative values were designated as zero. Data are represented as a percentage of total CD4+ or CD8+ T cells. (**A)** Pie graphs indicate the frequency of patients with (RESPONDER) or without (NON RESPONDER) detectable SARS-CoV-2 spike-specific CD4+ or CD8+ T cell responses, according to the different stages of disease (MMD n=7 and MMR n=9). **(B)** Percentage of SARS-CoV-2 spike-specific CD4+ T cells expressing IL-2, IFN-γ, or TNF-α and (**C**) percentage of SARS-CoV-2 spike-specific CD8+ T cells expressing IL-2, IFN-γ, or TNF-α in POST (orange ●) and BOOSTER (blue ▲) samples. Individual data points are shown, and lines connect the paired samples. Dotted lines show the lower limit of detection at <0.01. Statistical analysis was performed using a paired, two-tailed, nonparametric Wilcoxon test. *P* values shown when *P* < 0.05 (\**P* < 0.05, \*\**P* < 0.01).

We observed an increase in the percentage of MM patients with detectable SARS-CoV-2 spike-specific CD4+ T cells (at least one cytokine production) (100% of MMD and MMR patients) (Fig. 5A). Notably, we found a significant increase in IL-2^+^CD4^+^ T cells in the BOOSTER compared to the POST among MMD patients. MMR patients did not reach statistical significance as group, but 6/9 (66.6%) patients showed an increase in IL-2^+^CD4^+^ T cells. Intriguingly, the 4 MMR patients, that reached delectable levels of NAbs against Omicron variant after BOOSTER, had a concomitant increase in the percentage of IL-2^+^CD4^+^ T cells. In our cohort, we did not find any significant differences between BOOSTER and POST, in terms of IFN-γ^+^ or TNF-α^+^CD4^+^ T cells (Fig. 5B).

The percentages of patients with detectable SARS-CoV-2 spike-specific CD8^+^ T cell responses increased to 85.7% in MMD and, on the other hand, the percentages decreased to 55,5% in MMR patients (Fig. 5A). In fact, we found a significant reduction in SARS-CoV-2 spike-specific IFN-γ^+^CD8^+^ T cells in MMR patients in BOOSTER compared to POST samples (Fig. 5C), while we did not find any significant differences between BOOSTER and POST, in terms of IL-2^+^ or TNF-α^+^CD8^+^ T cells both in MMD and MMR patients. (Fig. 5C)

Taken together, these data demonstrate that booster immunization improved SARS-CoV-2 spike humoral and cellular responses in MMD patients and in most, but not all, MMR patients. After the booster dose, a significant increase of the NAb titers against almost all the analyzed variants was achieved in MMD but in MMR patients Omicron retain a negative impact.

## Discussion

In this study, we performed a comprehensive analysis of immune responses after mRNA SARS-CoV-2 vaccination among a cohort of patients with monoclonal gammopathies at different stages of disease, that are known to show a impaired or reduced response to vaccination.^5^ In addition, we also evaluated the impact of the different SARS-Cov-2 variants and the effect of booster dose on the immune response. Firstly, consistently with recent literature data, we found that MMR patients had significantly lower spike-IgG-Abs compared to the other groups of patients.^9,10,27^

Considering pre-vaccination immune parameters, immunoparesis with the involvement of more than one class of Ig, the reduction of absolute number of CD4^+^ T cells and consequently a reduced CD4^+^/CD8^+^ ratio may affect the production of spike-IgG-Abs after full vaccination. MM patients are known to have a deep reduction of CD4^+^ T cells with an inverted CD4^+^/CD8^+^ ratio compared to precursors stages of the disease.^28^ In fact, CD4^+^ T cells are closely associated with B cell differentiation into IgG-producing plasma cells and with the development of an adequate humoral immune response.

Only a fraction of the spike-IgG-Abs produced after vaccination are capable of neutralization, resulting in a blunted virus infection. We reported that anti-SARS-CoV-2 spike neutralizing titers for the original Wuhan-Hu-1 spike protein were strongly correlated with the spike IgG antibody levels. In particular, MMR patients showed a reduced NAb titers compared to all the other groups of patients, especially to MGUS, as previously reported.^10^ Several SARS-CoV-2 variants have emerged in the last year with an impact on vaccine efficacy^29^. Delta variant was dominating the pandemic at the time of vaccination of our patients, while the novel Omicron variant is currently dominating.^13^ It is reported in healthy subjects a reduction of NAb titers for Beta and Delta variants and an highly resistance of Omicron variant against antibody-mediated neutralization after both homologous and heterologous vaccination.^14,30,31^ Thus we have analyzed the susceptibility of five variants, including the Omicron one, to vaccine-induced antibody neutralization in our cohort of patients. In our study, after full vaccination, NAb titers against Omicron variant dramatically dropped compared to those against the original strain or the other four variants, with no MMR patients showing detectable levels of NAbs. Interestingly, all the analyzed variants, remarkably the Omicron one, had a significant negative impact on the neutralizing ability of the vaccine-induced antibodies not only in MMD and MMR but also in SMM, suggesting that this last group of patients already had features of immune system impairment more similar to MMD that MGUS patients. On the other hand, MGUS patients showed a lower neutralizing ability only to the Beta and Omicron variants, as reported in general population.^15,16,31,32^

Although NAbs are likely to be crucial in vaccine-induced protection, precise correlation to immunity are not completely defined and recent evidence also suggests an important role for T cells.^23,24,33,34^ Until now, only two groups described variable results on the antigen-specific cellular responses in the MM setting. In fact, Aleman et al. ^11^ highlighted that fully vaccinated seronegative MM patients had a reduction in the percentage of spike-specific CD4^+^ T cells compared to the seropositive patients. On the other hand, another study on MM patients showed a significant lower percentage of spike-specific CD8^+^INF-γ^+^ T cells in the seronegative group.^35^ Our study explores the vaccine-induced cellular responses also in the premalignant monoclonal gammopathies and our data indicate that 100% of the MGUS patients mounted a SARS-CoV-2 spike-specific CD4^+^ and CD8^+^ T cell response while the responder patients percentages decreased in MMD, MMR, and also SMM groups. In particular, a significant reduction of IL-2^+^CD4^+^ T cell percentage was described in MMD, MMR, and SMM compared to MGUS patients. It is reported that the development of antigen-specific IL-2^+^CD4^+^ T cells are essential for the differentiation of IgG-secreting plasma cells in humans.^36^ These data, together with the ones of the NAb titers, suggest that SMM patients have already dysregulated immune system which impairs both the humoral and CD4^+^ responses to vaccination.

Looking at CD8^+^ T cells, critical for the clearance of virus infected cells, MM patients, showed a reduced cytotoxic function exerted by spike-specific IFN-γ^+^ and TNF-α^+^ CD8^+^ T cells, compared to MGUS patients. These results could be related to a more severe breakthrough COVID-19 in vaccinated MM patients.

The decline of vaccine-induced immunity during time and the emergence of new variants led to the administration of a booster dose of SARS-CoV-2 vaccine. Our data demonstrate that booster immunization improved spike-specific humoral responses in MMD patients, as similar reported in a recent study^37^. Interestingly, a significant increase of the NAb titers against all the analyzed variants was achieved after the booster dose compared to full vaccination. The booster dose leads MMD patients to produce enough spike-IgG-Abs, and consequently spike-specific NAbs, that erased the negative impact of four spike variants seen after the full vaccination, but not of Omicron one.

On the other hand, MMR benefited from the booster dose, even if they maintained lower levels of spike-IgG-Abs and NAbs compared to MMD patients. In fact, a significant increase of NAb titers was observed against 3 out of 5 analyzed variants. A slightly increase was reported also against Omicron one, where almost half of MMR patient reach detectable levels of NAbs, but, overall, MMR patients retained an increased susceptibility to Beta, Delta and Omicron variants as reported after full vaccination.

Concerning the impact of the booster dose on antigen-specific T cells, interestingly, both MMD and MMR reached the 100% of responders in terms of SARS-CoV-2 spike-specific CD4^+^ T cells with an increased percentage of IL-2^+^CD4^+^ T cells compared to full vaccination. After booster dose, SARS-CoV-2 spike-specific CD8^+^ T cells had a highly variable trend in MM patients and, notably, almost all MMR patients had undetectable percentage of SARS-CoV-2 spike-specific IFN-γ^+^CD8^+^ T cells. This effect could be related to a different kinetic and/or homing of these circulating CD8^+^ T cells, as reported after COVID-19 in natural infection.^38^

Although our study analyzed a limited number of patients for each group of disease stage and it lacks healthy donors as controls, it is important to note that the groups are very homogenous for age and that all patients underwent the same vaccine administration scheme and blood collection time points, thus excluding potential bias related to age, vaccine type or timing of antigen-specific responses detection. A previous study ^10^, exploring the production of NAbs in a large cohort of vaccinated patients with plasma cell neoplasms, reported that patients with MGUS did not have significant differences compared to healthy controls, suggesting that these patients could be considered proper controls of an optimal response to SARS-CoV-2 vaccination.

In conclusion, our study indicates that patients with monoclonal gammopathies share a different and variable cellular and humoral immunity to SARS-CoV-2 vaccination and a significant reduced response was reported in MM patients with relapsed or refractory disease that underwent to at least two lines of treatment compared to MGUS patients. For the first time, our study underlines the negative impact of Omicron variants on the neutralizing ability of the vaccine-induced antibodies in MM and also in SMM patients after a full vaccination. Interestingly, the booster dose rescues the negative impact of spike variants on humoral response efficacy in all MMD patients and only partially in MMR patients, suggesting that this last group of patients needs to be considered at risk to develop a clinically relevant COVID-19 disease even after complete vaccination cycle. These data give the rational to carefully monitor vaccinated MM patients for SARS-CoV-2 infection and to consider further prophylactic approach, as the fourth vaccine dose, or preventive measures especially in patients under several lines of treatment.

## Data Availability

All data produced in the present study are available upon reasonable request to the authors

## Acknowledgements

This study was supported by funding from: AIRC under IG IG2017 ID. 20299 project and International Myeloma Society (IMS) under “Paula and Rodeger Riney Foundation Translational Research Grant” (PI Nicola Giuliani) and by the Associazione Italiana contro Leucemie, Linfomi e Mielomi ONLUS, ParmAIL.

## Author contributions

The study was conceptualized by N.G. and G.D.; P.S., V.M. and R.V. performed ELISA assay and PBMCs stimulation; G.D. prepared the pseudoviruses and performed the neutralization assays supported by V.F. and L.R.; R.V. performed the flow cytometry analysis; V.R., D.T., J.B.G, F.C. and N.T.I. processed the blood samples and collected sera and mononuclear cells; M.G., L.N., N.S., B.D.P. and N.G. provide clinical data and enrolled the patients; P.S., V.M. and R.V. analyzed data; P.S., R.V., G.D. and N.G. wrote the manuscript with input from all authors.

## Competing Interests

The authors have no relevant affiliations or financial involvement with any organization or entity with a financial interest in or financial conflicts with the subject matter or materials discussed in the manuscript apart from those disclosed.

## Notes

### Competing Interest Statement

The authors have declared no competing interest.

### Author Declarations

The study was approved by the Area Vasta Emilia Nord (AVEN) Ethics Committee

