## Supplemental methods for "Impact of Omicron variant on the response to SARS-CoV-2 mRNA Vaccination in multiple myeloma"

### SUPPLEMENTAL DATA

### SUPPLEMENTAL METHODS

**Sample processing.** Samples were processed within 4 h of the blood draw. All sera were heat-inactivated for 30 min at 56°c before use in the assays. PB mononuclear cells (PBMCs) were obtained by density gradient centrifugation. Cells were resuspended in complete RPMI media supplemented with 10% heat-inactivated fetal calf serum (Biochrom, GmbH), 2 mM L-glutamine and 1% penicillin–streptomycin (R10) for counting and cryopreservation in cold freezing media (FCS containing 10% dimethylsulphoxide (DMSO)) at a concentration of 8–12 × $10^6$ cells per ml.

**Peptides and stimulations.** Peptides spanning the full length of the SARS-CoV-2 spike protein sequence were used in the antigen-specific T cell assay (PepMIX SARS-CoV-2 spikeglicoprotein, cat. PM-WCPV-S-3, JPT Peptide Techonologies GmbH). A total of 315 peptides, synthesized as 15-mers overlapping by 11 amino acids, were available in two separate peptide pools spanning the S1 (158 peptides) and S2 (157 peptides) subunits of the SARS-CoV-2 spike protein (original Wuhan-Hu-1 sequence). The freeze-dried pools were dissolved in DMSO, and stock aliquots were stored at -20°C until use.

**SARS-CoV-2 pseudoviruses generation and neutralization assay against the original viral strain and variants.** HEK 293T cells were transfected, in T175 cm2 flasks, with 25 µg of pLV-EF1α-(turboGFP-Luc2)-WPRE transfer vector, 15 µg of p8.74 packaging vector, 13 µg of pseudotyping vector coding for 6 different spike glycoproteins: Wuhan-Hu-1 (B.1 Lineage; China) Alpha (B.1.1.7. Lineage; United Kingdom), Beta (B.1.351 Lineage; South Africa), Gamma (P.1 Lineage; Brasil), Delta (B.1.617.2 Lineage, India) or Omicron (B.1.1.529 Lineage; Europe) (Supplementary Data 1) and 5 µg of pREV (58 µg of total DNA) diluted in 3 mL of complete DMEM (Euroclone) without serum and 145 µL of PEI (Polysciences, Inc., Warrington, PA, USA) (1 mg/mL in PBS) (ratio 1:2.5 DNA/PEI). After at least 15 min incubation at room temperature, 4× volumes of complete DMEM without serum were added, and the transfection solution was transferred to the cell monolayer. After 6 h of incubation at 37 °C and 5% CO2, in a humidified incubator, the transfection mixture was replaced with 25 mL of fresh complete EMEM supplemented with 10% FBS and incubated for 48 h at 37 °C and 5% CO2. The flask was then frozen–thawed at −80 °C; transfected cell supernatant (TCS) containing spike pseudovirus was clarified via centrifugation at 3500 rpm for 5 min at 4 °C, filtered through a 0.45 μm filter (Millipore, Merk, Darmstadt, Germany), aliquoted, tittered by limited dilution, and stored at −80 °C. 25μL of complete medium are added to each well of the microplate and 25 μL of each serum is added to each well of the first line of wells. Then 25 μL of the first dilution of the sera (1:2) are mixed and passed to the subsequent lines of wells. 25μL from the last line of wells are discarded. Therefore, the final volume for each well is 25μL. Twenty-five microliters of spike pseudovirus preparation diluted in complete EMEM with 10% FBS (corresponding to ~104 relative luciferase units (RLUs); ~3–5μL of the initial preparation) was added to each well and left to incubate at room temperature for 1.5 h. Final volume for each well reached 50 μL; therefore, the sera dilution was doubled, 1:4–1:8–1:16–1:32–1:64–1:128–1:256–1:512. Next, 50 μL of complete EMEM with 10% FBS, containing 104 HEK/ACE2/TMPRRS2/Puro cells [20], was added to each well and left for 60 h at 37 °C and 5% CO2. Plates were read by adding 25 μL of complete EMEM containing luciferin to each well just before the reading of the microplate with the luminometer (Victor, Perkin Elmer).

**Intracellular Cytokine Staining Flow Cytometry (ICS) T Cell Assay Quality**

BDCompBeads (catalog no. 552843) and PBMCs were used for single-fluorochrome compensation. Experiments were performed with protocols optimized to reduce batch variation and to ensure mixing of patient groups across batches. BD FACSDiva CS&T research beads (catalog no. 655050) were used to track and adjust photomultiplier tubes over time. Paired samples from PRE and POST vaccination of each patients were ran in parallel in 13 independent batches. BOOSTER samples were ran in 3 independent batches. To maintain reproducibility between sample batches, the same lot of antibodies were used for all samples and a single operator performed gating and analysis. All FACS plots were manually inspected to confirm quality of staining and cytometry acquisition.

**Statistical Analysis**

Unpaired samples were compared using Kruskal-Wallis and Mann–Whitney U tests, and paired samples were compared with the Wilcoxon test. Correlation coefficients were quantified by the Spearman rank correlation coefficient. All tests were performed in a two-sided manner, using a nominal significance threshold of $P < 0.05$ unless otherwise specified.
