## Supplemental table 1 for "Impact of Omicron variant on the response to SARS-CoV-2 mRNA Vaccination in multiple myeloma"

**Supplemental Table 1: Patients' Clinical Data**

| | MGUS (n=6) | | SMM (n=10) | | MMD (n=9) | | MMR (n=13) | |
|---|---|---|---|---|---|---|---|---|
| Age (year) | 74 | [61-84] | 77 | [51-86] | 69 | [51-83] | 80 | [63-83] |
| Female gender | 100% | (6) | 40% | (4) | 16,60% | (2) | 53,80% | (7) |
| Years from diagnosis | 7 | [3-15] | 9,5 | [3-15] | 2 | [1-6] | 6 | [2-10] |
| Lymphocyte Absolute counts ($x10^{-3}/\mu l$) | 2,07 | [1,6-4,04] | 1,44 | [0,67-3,85] | 1,2 | [0,54-2,09] | 0,97 | [0,4-3,17] |
| **Immunoparesis** | | | | | | | | |
| None | 16,6% | (1) | 20% | (2) | 0% | (0) | 0% | (0) |
| 1 class | 66,7% | (4) | 20% | (2) | 66,7% | (6) | 23,1% | (3) |
| 2-3 classes | 16,6% | (1) | 60% | (6) | 33,3% | (3) | 76,9% | (10) |
| **ISS** | | | | | | | | |
| I | | | | | 33,3% | (3) | 23,1% | (3) |
| II | | | | | 55,5% | (5) | 30,8% | (4) |
| III | | | | | 11,1% | (1) | 46,1% | (6) |
| **Mayo Score** | | | | | | | | |
| 0 | 0,0% | (0) | | | | | | |
| 1 | 33,7% | (2) | | | | | | |
| 2 | 66,7% | (4) | | | | | | |
| 3 | 0,0% | (0) | | | | | | |
| **2-20-20 Score** | | | | | | | | |
| 0 | | | 10,0% | (1) | | | | |
| 1 | | | 50,0% | (5) | | | | |
| 2-3 | | | 40,0% | (4) | | | | |
| **Previous line of therapy (n)** | | | | | 1 | [1-2] | 2 | [2-6] |
| **Disease reponse status:** | | | | | | | | |
| CR or sCR | | | | | 33,3% | (3) | 46,1% | (6) |
| VGPR or PR | | | | | 55,5% | (5) | 46,1% | (6) |
| SD or PD | | | | | 11,1% | (1) | 7,8% | (1) |
| **Treatment regimen contains:** | | | | | | | | |
| IMiDs | | | | | 55,5% | (5) | 61,5% | (8) |
| PI | | | | | 22,2% | (2) | 7,8% | (1) |
| Steroids | | | | | 44,4% | (4) | 69,2% | (9) |
| Anti-CD38 mAb | | | | | 0,0% | (0) | 30,8% | (4) |
| No active tratment | | | | | 22,2% | (2) | 23,1% | (3) |

**Note**: values are presented as percentage (n) ore median [range]

**Abbreviations**: MGUS: Monoclonal Gammopathy of Undetermined Significance; SMM: Smoldering Multiple Myeloma; MMD: Multiple Myeloma Newly Diagnosed; MMR: Multiple Myeloma Refractory/Relapsed; ISS: International Staging System; CR: Complete Response; sCR: stringent Complete Response; VGPR: Very Good Partial Response; PR: Partial Response; SD: Stable Disease; PD: Progressive Disease; IMiDs: Immunomodulatory Drugs; PI: Proteasome Inhibitors; n= number
