## Supplemental figure for "Impact of Omicron variant on the response to SARS-CoV-2 mRNA Vaccination in multiple myeloma"

**Supplemental Figure 1**

**A**

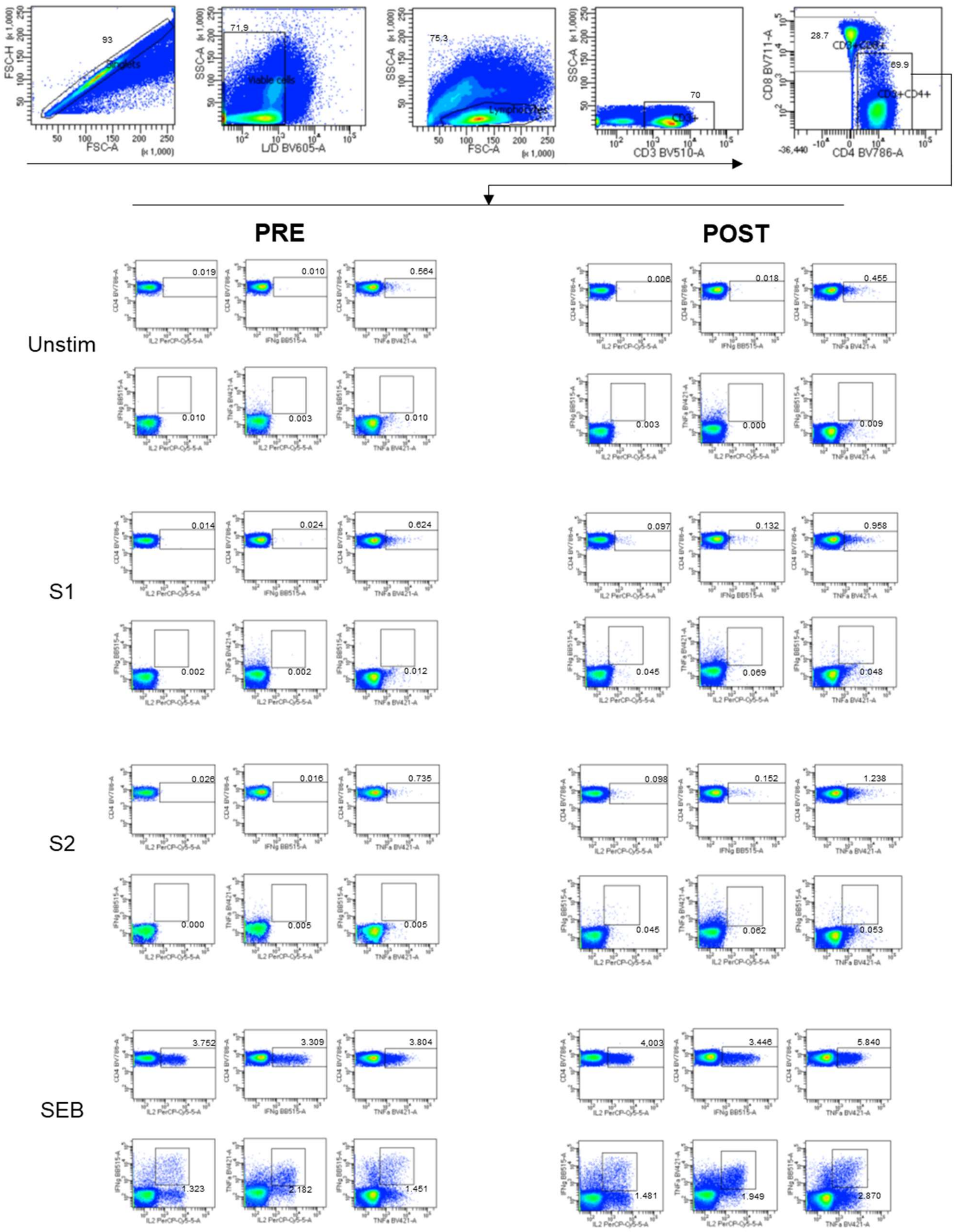

**B**

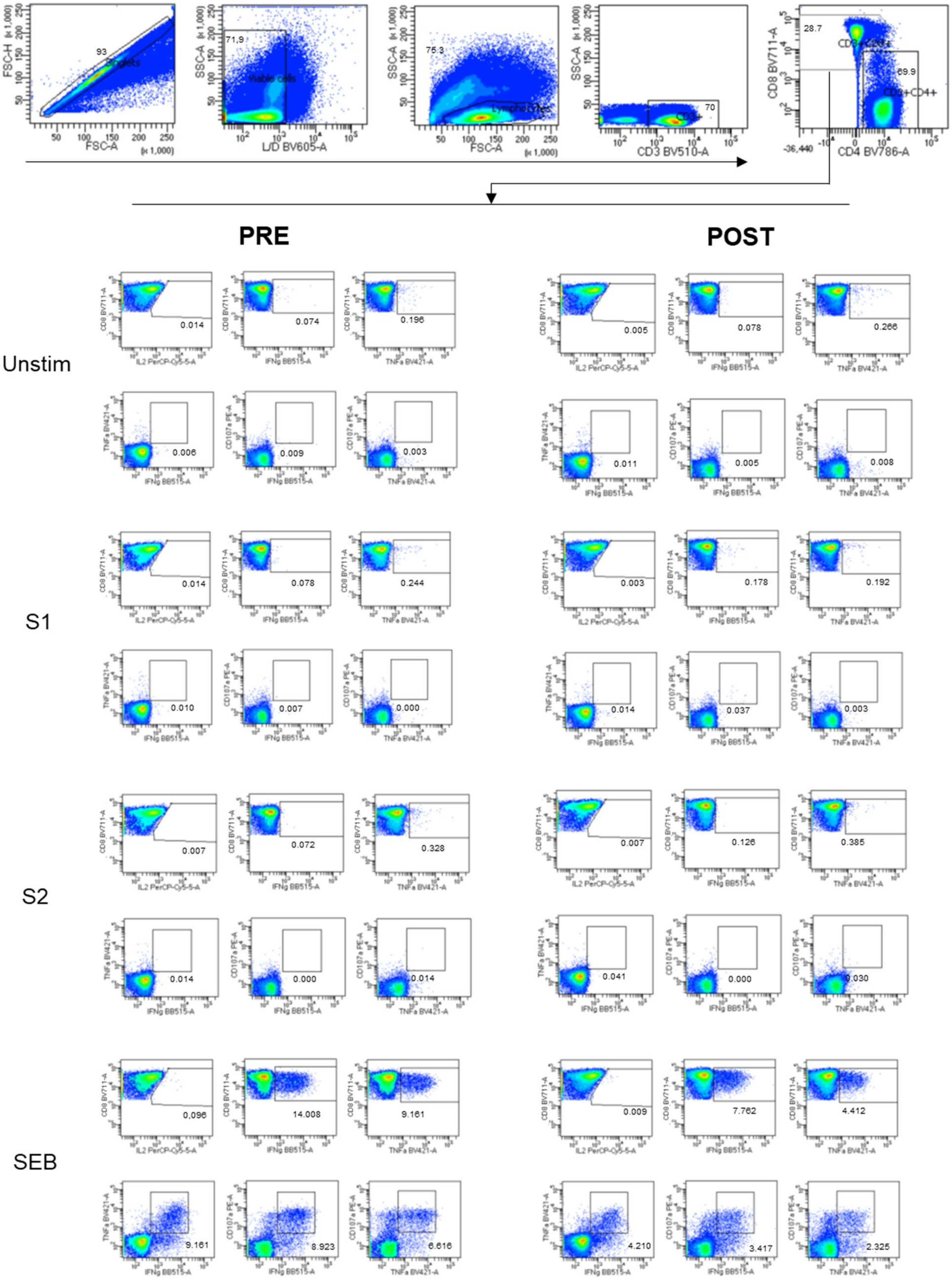


**Supplementary Figure 1.** Exemplar gating strategy for intracellular cytokine staining from a representative patient sample. (**A**) Parent gating strategy for selection of viable CD3+CD4+

lymphocytes and gating for quantification of cytokine positive CD4+ T cells. Unstim = unstimulated sample, S1 = sample stimulated with peptide pool spanning the S1 subunit of the SARS-CoV-2 spike protein, S2 = sample stimulated with peptide pool spanning the S2 subunit of the SARS-CoV-2 spike protein, SEB = sample stimulated with *S. enterotoxin* B. Background subtraction (S1- unstim and S2- unstim) was calculated following gating**. (B**) Parent gating strategy for selection of viable CD3+CD8+ lymphocytes and gating for quantification of cytokine and CD107a positive CD8+ T cells. Unstim = unstimulated sample, S1 = sample stimulated with peptide pool spanning the S1 subunit of the SARS-CoV-2 spike protein, S2 = sample stimulated with peptide pool spanning the S2 subunit of the SARS-CoV-2 spike protein, SEB = sample stimulated with *S. enterotoxin* B. Background subtraction (S1- unstim and S2- unstim) was calculated following gating.

**Supplemental Figure 2**

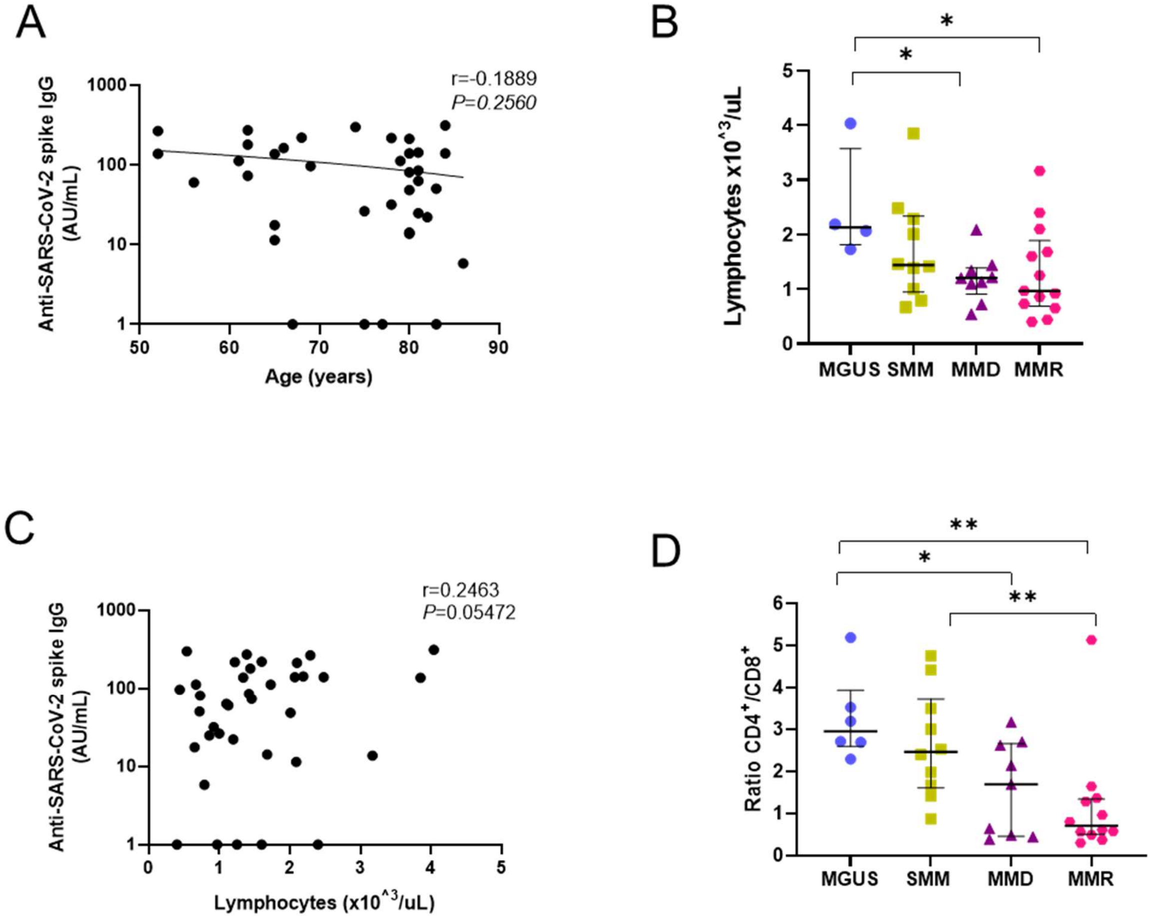


<u>**Supplemental Figure 2.**</u> <u>**(A)**</u> Correlations between levels of SARS-CoV-2 spike IgG antibodies after vaccination and age in years of the patients in the cohort. Correlations were calculated using nonparametric Spearman rank correlation. (**B)** Baseline (PRE) absolute number of lymphocytes in PB blood in patients subdivided in the different stages of disease: MGUS = indigo circle (n =6); SMM = avocado square (n =10); MM = purple triangle (n = 9) and MMR = fuchsia rhombus (n = 13). Significant difference was determined by two-tailed Mann–Whitney U tests. **(C)** Correlations between levels of SARS-CoV-2 spike IgG antibodies after vaccination and the baseline (PRE) absolute number of lymphocytes in PB blood were calculated using nonparametric Spearman rank correlation. (**D)** Baseline (PRE) RATIO $CD4^{+}/CD8^{+}$ cells in PB blood in patients subdivided in the different stages of disease: MGUS = indigo circle (n =6); SMM = avocado square (n =10); MM = purple triangle (n = 9) and MMR = fuchsia rhombus (n = 13). Significant difference was determined by two-tailed Mann–Whitney U tests. *P* values are shown when $P < 0.05$ (*$P < 0.05$, **$P < 0.01$).

**Supplemental Figure 3**

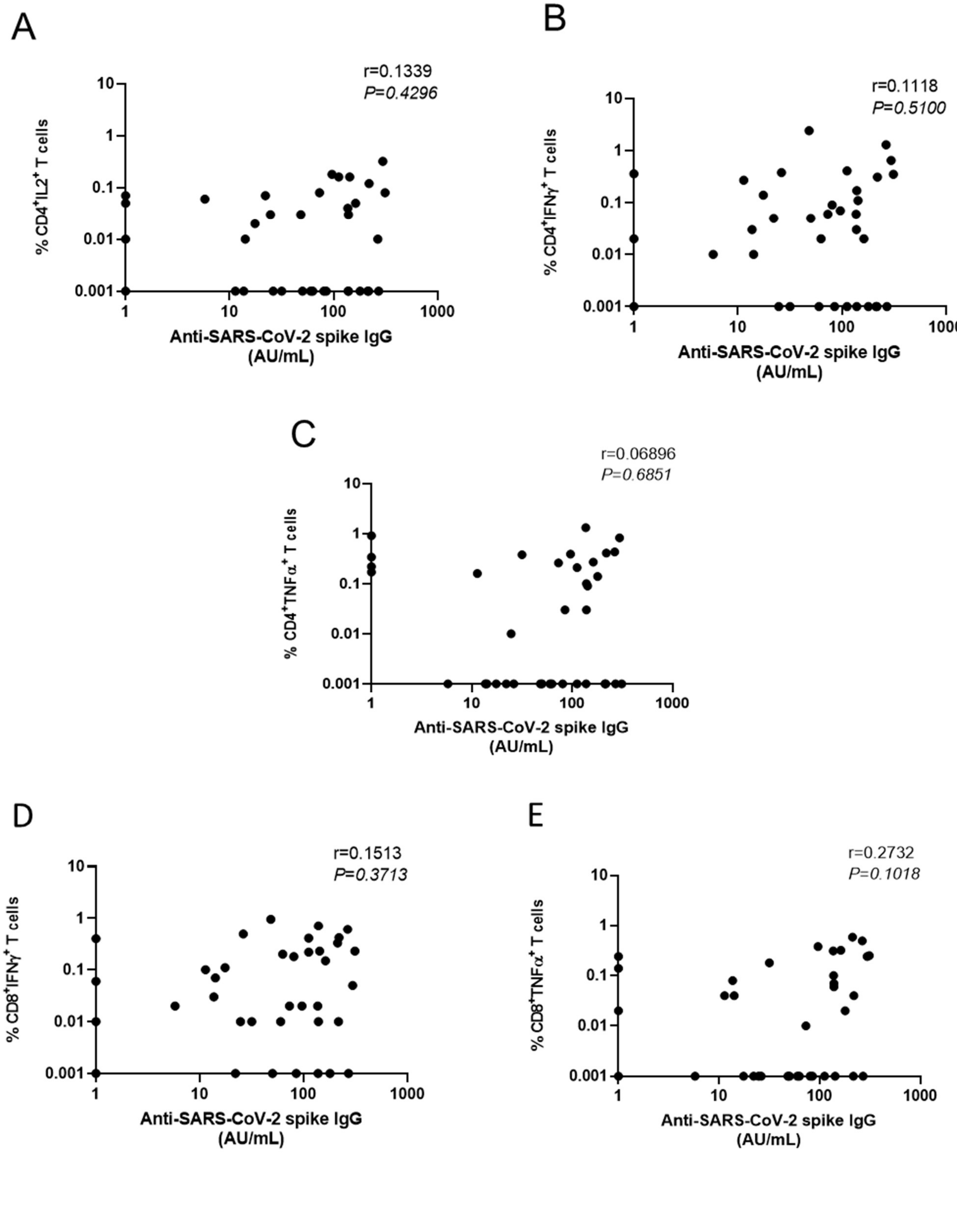


**Supplemental Figure 3.** Correlations between levels of SARS-CoV-2 spike IgG antibodies after vaccination and percentage of SARS-CoV-2 spike-specific CD4$^+$ T cells expressing (**A**) IL-2, (**B**) IFN-γ, or (**C**) TNF-α and percentage of SARS-CoV-2 spike-specific CD8$^+$ T cells expressing (**D**) IFN-γ, or (**E**) TNF-α after vaccination. Correlations were calculated using non-parametric Spearman rank correlation.
